# Pre-COVID-19 *ex vivo* cross-reactive IFN-γ cellular response to SARS-CoV-2 spike overlapping peptides is more robust among Kenyan compared to Swedish adults

**DOI:** 10.1101/2025.01.24.25321024

**Authors:** Perpetual Wanjiku, Benedict Orindi, Jedidah Mwacharo, James Chemweno, Henry Karanja, Barbara Kronsteiner, Oscar Kai, Daniel Wright, Lynette Isabella Ochola-Oyier, Christopher Sundling, Susanna Dunachie, George M Warimwe, Anna Färnert, Philip Bejon, Francis M. Ndungu, Eunice Nduati

## Abstract

**Introduction:** Global WHO data indicate that Sub-Saharan African (SSA) countries, such as Kenya, experienced reduced coronavirus disease 2019 (COVID-19) severe-morbidity and mortality burdens relative to their more affluent counterparts in Europe, Asia, and North America.

**Methods:** We analysed peripheral blood mononuclear cells (PBMC) samples collected from Kenya and Sweden before and during COVID-19. Pre-COVID-19 samples were available for 80 adults and 10 infants from Kenya, and 20 adults from Sweden. COVID-19 samples were available for 39 Kenyan adults. The samples were analysed for *ex vivo* IFN-γ secretion using an Enzyme-Linked Immunosorbent (ELISpot) assay following *in vitro* stimulations with overlapping SARS-CoV-2 spike-protein peptides. T-cells expressing IFN-γ, IL-2, TNF-α, CD154, and CD107a were assessed following similar stimulations, using intracellular cytokine staining (ICS) and multiparameter flow cytometry.

**Results:** 55.7% of the Kenyan pre-COVID-19 adults were classified as responders by ELISPOT responses to spike-protein peptides, compared with 28% of Swedish pre-COVID-19 adults (p = 0.04). The frequencies for SARS-CoV-2 spike-specific TNF-α CD4+, TNF-α CD8+ and IFN-γ CD8+ T-cell responses, tended to be higher in the Kenyan adults although these differences did not reach statistical significance.

**Conclusion:** Pre-COVID-19 T-cell responses could contribute to lower morbidity and mortality associated with SARS-CoV-2 infections in SSA relative to Europe, Asia, and North America.

**Highlights:**

- Higher frequencies of cross-reactive IFN-γ-secreting cells among Kenyan pre-COVID-19 adults compared to Sweden.
- Trends of higher TNF-α and IFN-γ T-cell responses among Kenyan adults.
- Presence of pre-existing T-cell immunity in Sub-Saharan Africa.

## Introduction

Most SARS-CoV-2 infections result in either asymptomatic or mild disease, but a small proportion of individuals develop severe COVID-19 requiring hospitalisation, and some of these cases result in death (1). Compared to North America, Asia and Europe, SSA experienced relatively fewer severe cases and fatalities despite many reports of similar SARS-CoV-2-infection rates (2). This paradoxical observation has puzzled many public health experts (3–5). The suggested hypotheses for the reduced burden of severe and fatal COVID-19 in SSA include potential underreporting of cases and deaths, a relatively younger population, warmer climatic conditions, and immune regulation induced from either BCG vaccinations (6–8) or previous exposures to highly inflammatory diseases like malaria (9), or cross protection from a higher level of exposure to other coronaviruses (10–16).

There is evidence for cellular and humoral cross-reactivity to SARS-CoV-2 (10–16), and the role for pre-existing hCoV reactive CD4+ T-cell responses in providing SARS-CoV-2 cross protection (17). A high incidence of exposures to hCoVs and other pathogens could have induced pre-COVID-19 cross protective T-cell responses, or could have downregulated inflammation after infections of SSA residents with SARS-CoV-2 (11–16). SARS-CoV-2 cross neutralising antibodies are undetectable, even in recent confirmed cases of hCoV (18). However, antigen-specific memory T-cells are more durable and frequently cross-reactive, and may play a key role in the clearance of SARS-CoV-2 infected cells (13,19,20). Recent studies, predominantly conducted in high-income countries (HICs), have reported the presence of pre-COVID-19 cross-reactive T-cell immune responses to SARS-CoV-2 with endemic hCoVs (11–15,20). Studies from SSA countries like Uganda (21,22), Senegal (23), and South Africa (24) have provided some evidence regarding pre-COVID-19 cross-reactive T-cell to SARS-CoV-2 in adults.

This study examined *ex vivo* pre-COVID-19 IFN-γ cellular responses using ELISpot assay and the frequency of functional T-cells, expressing either IFN-γ, IL-2, TNF-α, CD154, and CD107a by multiparametric flow cytometry following *in vitro* stimulation with overlapping SARS-CoV-2 spike peptides. Importantly, we compared results from PBMC samples collected before 2019 from Kenyan and Swedish adults, undertaking standardised assays as a single batch in the same laboratory in Kenya. We further included additional control samples from 10 Kenyan infants (<1 year) and 39 adults COVID-19 patients.

## Methodology

### Study design and participants

Kenyan pre-COVID-19 blood samples were collected from 80 adults and 10 infants aged below 1 year during an annual malaria surveillance cross-sectional survey conducted in March 2018 in Kilifi. The Swedish pre-COVID-19 blood samples had been collected from 20 healthy adult volunteers living in Sweden between March 2012 and June 2019. At the time of pre-COVID-19 sampling, all the individuals had no symptomatic or acute infection. Post-COVID-19 positive control samples were collected from 39 Kenyan adults for studies investigating the kinetics of the immune response to SARS-CoV-2 in COVID-19 patients in Kilifi and Nairobi between 1^st^ August 2020 and 15^th^ February 2022. Blood was separated into plasma and PBMCs and stored at -80°C and liquid nitrogen, respectively, until used for analyses. All the participants consented for their respective samples to be used for both the contemporaneous and future research. This project received ethical approval from the Kenya Medical Research Institute’s (KEMRI) Scientific and Ethics Review Unit (SERU) under protocols SERU 3149, 4081 and 4085. Ethical approval for the collection of the Swedish samples was obtained from the regional Ethics Committee in Stockholm, Sweden under protocols 2006/893-31/4 and 2019-03436.

### Overlapping SARS-CoV-2 spike peptides and in vitro stimulants for PBMC in the ELISpot assay

Pools of peptides covering the spike region 1 (S1) and spike region 2 (S2) of the SARS-CoV-2 spike protein were synthesised by Mimotopes Pty Ltd. Each peptide was 15-mers long and overlapped with the next by 10 amino acid residues. A total of 12 mini-pools, P1 to P12 (Pro-immune), which corresponded to SARS-CoV-2 spike regions S1 (P1 to P6) and S2 (P7 to P12) (25) were used. Two positive controls, phytohemagglutinin-L (PHA-L, Sigma) and anti-CD3, were used to induce a mitogenic response and demonstrate viability of lymphocytes and T-cells, respectively. A negative control (i.e., no peptide but with the Dimethyl sulfoxide [DMSO] carrier) was also included to detect non-specific IFN-γ release. The first 10 samples with sufficient cell numbers to test against all 12 peptide pools were included in a preliminary analysis to guide peptide pool prioritisation when participant’s PBMCs recovered after thawing yielded insufficient numbers for testing across all pools. Following this analysis, 4 peptide pools, 3 and 4 representing S1 region, and 8 and10 representing S2 region, were prioritised when the numbers of PBMCs were inadequate (Figure S1).

### ELISpot assay

We used an IFN-γ ELISpot assay, which quantifies the frequency of antigen-specific effector cells producing IFN-γ after an overnight co-culture of PBMC with either specific peptides or a non-antigen specific positive control (25). Briefly, 96 well ELISpot plates (Millipore, UK) were coated with a monoclonal antibody against IFN-γ (Mabtech, AB, Sweden) at 10µg/mL overnight. 200000 PBMCs per well were plated in triplicates for each individual and plates incubated at 37°C in a humidified carbon dioxide (5%) incubator for 16–18 hours of stimulation with overlapping peptide pools at 10µg/mL in triplicate wells. After incubation, PBMCs were discarded, the plate washed, and the detector for the secreted IFN-γ footprints on the plate membrane, anti-IFN-γ biotinylated monoclonal antibody (Mabtech), added at 1µg/mL and the plate incubated for 2 hours at room temperature. The plate was then washed, and streptavidin alkaline phosphatase (Mabtech) added at 1µg/mL, before another incubation for 1 hour at room temperature. Following washing, the 1-StepTM NBT/BCI (nitro blue tetrazolium/5-bromo-4-chloro-3-phosphatase) substrate (Thermo Scientific) was added, and the plate left in the dark for 7 minutes until the developing spots were clearly visible. The plate was then washed three times using tap water. Following overnight drying, the spots in each well were enumerated with an AID ELISpot Reader (v.4.0). Counts were presented as IFN-γ Spot-Forming Units (SFUs) per million PBMC, which is determined by taking an average of the triplicate wells. Results are reported here as SFUs per million PBMCs after subtracting the background (mean of individual negative wells) from the antigen-specific responses. We summed up the responses from pools P3 and P4 (representing S1) and pools P8 and P10 (representing S2). For the spike region, we combined the aggregated responses from S1 and S2. Individual’s assays were considered failed, and their results excluded if the negative control wells had >150 SFUs per million PBMCs, or the positive control wells had <200 SFUs per million PBMCs. An ELISpot assay limit of detection of 10 SFUs per million PBMCs was also applied, and all wells with values <10 SFUs per million PBMCs replaced with a possible minimal result equal to 5 SFUs per million PBMCs. Individuals’ IFN-γ cellular responses were considered positive, if their SFUs were >71 SFUs per million PBMCs after background subtraction (corresponding to the 25^th^ percentile of Kenyan COVID-19 patients).

In total, 79 of 80 pre-COVID-19 Kenyan adults, 10 of 10 of the Kenyan infants, 18 of 20 Swedish adults and 37 of 39 post-COVID-19 Kenyan adult patients diagnosed with COVID-19 were included in the analysis. Of participants included in the analysis, 78 (99%) pre-COVID-19 Kenyan adults, 3 (30%) Kenyan infants, 5 (28%) Swedish adults and 14 (38%) COVID positive Kenyan patients had all the 12 peptide pools tested (Table S1). Due to the small number of infants, Swedish adults and COVID cases with entire 12 peptide pools tested, subsequent analyses were restricted to the prioritised pools 3 and 4 for S1 responses and pools 8 and 10 for S2 responses, and a summation of pools 3, 4, 8 and 10 for total spike responses (Figure S1).

### Synthetic peptide pools for intracellular cytokine staining stimulation assay

A total of 178 peptides (15–18-mers with a ten amino acid overlap) spanning the entire SARS-CoV-2 spike protein (S: positions 1–178) were synthesised by Mimotopes Pty Ltd and pooled together into one pool for PBMC stimulations. DMSO was used as a negative control and PMA/ionomycin as a positive control.

### Intracellular cytokine staining and multi-parametric flow cytometry

To determine T-cell function, we used ICS and multiparametric flow cytometry to identify and quantify effector T-cells responding to *in vitro* stimulation by overlapping SARS-CoV-2 spike peptides with IL-2, IFN-γ and TNF-α cytokine secretion, and upregulation of CD154 and CD107a activation makers. Briefly, cryopreserved PBMCs were thawed and rested for 2– 4 hours and thereafter plated at 1 × 10^6 live cells per well for SARS-CoV-2 spike peptide stimulations and the DMSO, and at 0.5 × 10^6 live cells per well for PMA/ionomycin in 96-well round-bottom plates. SARS-CoV-2 spike peptide stimulation was done at a final concentration of 2 µg/mL, PMA at 0.05 µg/mL and ionomycin (Sigma) at 0.5 µg/mL, and DMSO (Sigma) at 2 µg/mL. All the stimulations and controls were done in the presence of the co-stimulatory anti-CD28 and anti-CD49 (1 µg/mL, BD Biosciences) antibodies. CD107a BV421 (0.04 μg/ml, BD Biosciences) was added to all wells and cells incubated for 1 hour at 37°C temperature, 5% CO_2_, 95% humidity for surface staining. Following incubation, Brefeldin A (5 µg/mL, MP Biomedicals) was added, and cells incubated for a further 15 hours. After washing the cells with 1X PBS, dead cells were labelled using live/dead fixable aqua dye (Invitrogen) for 30 mins at 25°C. The cells were then washed with 1X Perm/Wash before fixation and permeabilisation using Cytofix-Cytoperm for 20 mins at 4°C. Cells were washed three times with 1X Perm/Wash and then stained with APC ‘Fire’ 750 anti-CD3 (Biolegend), PE Dazzle 594-anti-CD4 (Biolegend), PercpCy5.5-anti-CD8 (Biolegend), PECy7-anti-CD154 (Biolegend), APC-anti-IFN-γ (BD Biosciences), PE-anti-IL2 (Biolegend) and FITC-anti-TNF-α (BD Biosciences) for 20 mins at 4°C. Cells were washed twice with 1X Perm/Wash and resuspended in 200 μl 1X PBS and stored at 4°C in the dark until acquisition. Samples were acquired on a BD LSR Fortessa (BD Biosciences) flow cytometer, and data analysed using FlowJo version 10.10.0 software. Gating strategies used are detailed in (Figure S2). Results are presented as the proportion of IFN-γ, IL-2 or TNF-α secreting CD4 or CD8 T-cells, or as the proportion of CD154 expressing CD4 T-cells or CD107a expressing CD8 T-cells after subtracting the background (values from negative control wells for each sample). A limit of detection of 0.02% was applied, and all wells with values below 0.02% were replaced with the lowest possible value of 0.01%. Boolean gates were used to examine the frequency of polyfunctional T-cells defined as CD4 or CD8 T-cells co-expressing any of the cytokines IFN-γ, TNF-α, IL-2 and surface markers CD154 or CD107a.

### Statistical analysis

Data management and statistical analysis were performed using GraphPad Prism version 10.2.3 (26). IFN-γ SFUs detected by ELISPOT were log-transformed prior to analysis. For the primary analysis, pairwise comparisons were performed between Kilifi pre-COVID-19 adults and Swedish pre-COVID-19 adults to assess differences in IFN-γ SFUs, as well as the proportions of cytokine secreting T-cells and T-cell activation markers in response to SARS-CoV-2 spike peptides or its subregions (S1 or S2). A two-sample t-test was used to compare IFN-γ SFUs as continuous variables, while Fisher’s exact test was applied to compare the proportions of responders to SARS-CoV-2 spike peptides between Kilifi pre-COVID-19 adults and Swedish pre-COVID-19 adults. The Mann Whitney test was used for comparisons of the proportions of cytokine-secreting T-cells and T-cell activation markers. For the secondary analysis, a one-way analysis of variance (ANOVA) with Tukey’s multiple comparisons test was used to evaluate differences in IFN-γ SFUs across all four study groups. The Kruskal Wallis test with Dunn’s multiple comparisons test was used to compare the proportions of cytokine secreting T-cells and T-cell activation markers in response to SARS-CoV-2 spike peptides or subregions across the study groups. A paired t-test was used to assess interindividual variability in responses to peptides corresponding to S1 and S2. All tests were performed at 5% significance level.

## Results

### Participants characteristics

The median ages of the pre-COVID-19 study participants were 31 years (IQR 24-40 – for Kenyan adults, 33 years (IQR 31-41) for Swedish adults, and 0.75 years (IQR 0.58-0.83) for Kenyan infants. The proportions of male participants were 29% for Kenya adults, 5% for Swedish adults and 50% for Kenyan infants. Kenyan COVID-19 patients had a median age of 43 years (IQR 34-55), with about one in two being male (Table S2).

### IFN-γ responses by ELISpot

For the S1 region, Kenyan pre-COVID-19 adults tended to have higher IFN-γ responses (with a geometric mean of 42 [95% CI 31–56] SFUs per million PBMCs) than Swedish pre-COVID adults (geometric mean of 22 [95% CI 12–39] SFUs per million PBMCs; p = 0.05). For the S2 region, Kenyan and Swedish pre-COVID-19 adults had similar geometric mean responding T-cell numbers at geometric mean of 24 (95% CI: 19–30) and 27 (95% CI: 16– 45), SFUs per million PBMCs, respectively (*p* =0.66; Figure 1a).

**Figure 1.**
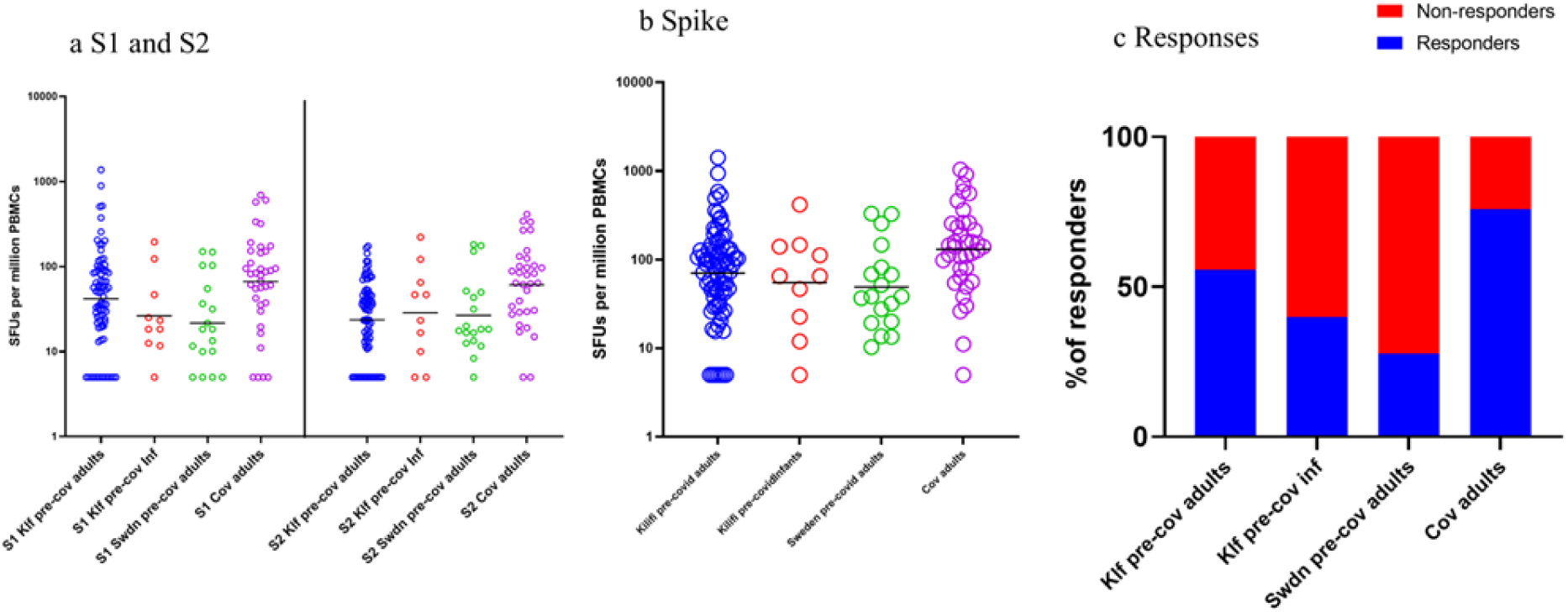
Comparison of the frequencies of IFN-gamma secreting cell responses to SARS-CoV-2 Spike regions 1 and 2 among study groups. **a** frequencies of Spike regions 1 and 2 separately, **b** aggregated frequencies of Spike regions 1 and 2, **c** Responders (Cut off for responders is 25^th^ percentile of Spike in CoV-adults). Comparisons done using t-test, ordinary one-way ANOVA with Tukey’s multiple comparisons test and Fisher’s exact test. Black line shows Geometric mean.

With S1 and S2 regions combined, Kenyan pre-COVID-19 adults had higher responses (geometric mean of 70 [95% CI 53–93]) than Swedish pre-COVID-19 adults (49 [95% CI 29–84]; p = 0.26; Figure 1b). Amongst the Kilifi pre-COVID-19 adults there were higher S1 responses compared to S2 responses (geometric mean of 42 [95% CI 31–56] vs. 24 [95% CI 19–30] SFUs per million PBMCs; p < 0.0001) (Figure S3a). For the Swedish pre-COVID-19 and infants pre-COVID-19, the magnitude of S1 and S2 responses were similar (Figures S3a– c).

We next analysed the proportion of responders within each subgroup. Responders were defined as individuals with IFN-γ SFUs responses exceeding 71 SFU per million PBMCs after background subtraction, corresponding to the 25th percentile of Kenyan COVID-19 patients. Notably, 55.7% of the Kenyan pre-COVID-19 adults were classified as responders, which was significantly higher than 28% of Swedish pre-COVID-19 adults (p = 0.04) in response to SARS-CoV-2 spike peptides. Among Kenyan pre-COVID-19 infants, 40% were responders. As expected, a large proportion, 75.7% of the COVID-19 positive individuals were classified as responders (Figure 1c).

### Cross reactive cytokine responses to SARS-CoV-2 peptides by ICS

To establish whether the IFN-γ responses observed by ELISpot were from memory T-cells and whether other cytokines were secreted, we incorporated ICS assay followed by multiparametric flow cytometry. Our data show that both CD4+ and CD8+ T-cells from the different groups produced IFN-γ, TNF-α and IL-2 after incubation with overlapping SARS-CoV-2 spike peptides. CD4+ T-cells also upregulated CD154 an activation marker. Kenyan pre-COVID-19 adults exhibited higher cytokines levels for IFN-γ and TNF-α responses, though not statistically significant (Figure 2). For the IFN-γ CD8+ T-cell responses, Kenyan pre-COVID-19 adults had a higher median of 0.06% (IQR 0.01–0.54%) compared to Swedish pre-COVID-19 adults (median: 0.01% [IQR 0.01–0.4%]; p = 0.54). The TNF-α CD4+ T-cell responses were higher in Kenyan pre-COVID-19 adults (median: 0.03% [IQR 0.01–0.095%]) than Swedish pre-COVID-19 adults (median: 0.015% [IQR 0.01–0.055%]; p = 0.39). Similarly, TNF-α CD8+ T-cell responses were elevated in Kenyan pre-COVID-19 adults (median: 0.06% [IQR 0.01–0.17%]) compared to Swedish pre-COVID-19 adults (median: 0.03% [IQR 0.01–0.05%]; p = 0.13) (Figure 2b, c–d). The Kenyan COVID-19 patients had significantly higher TNF-α CD4+ T-cells levels (median: 0.26% [IQR 0.11– 0.73%]) compared to Kenyan pre-COVID-19 adults, (median: 0.03% [IQR 0.01–0.095%]; p=0.01), Kenyan pre-COVID-19 infants (median: 0.02% [IQR 0.01–0.04%]; p= 0.007) and Swedish pre-COVID-19 adults (median: 0.015% [IQR 0.01–0.055%]; p= 0.005) (Figure 2c). IL-2 CD4+ T-cell frequencies were also higher in Kenyan COVID-19 patients (median: 0.09% [IQR 0.015–0.515%]) compared to Kenyan pre-COVID-19 adults (median: 0.01% [IQR 0.01–0.025%]; p= 0.02) and Swedish pre-COVID-19 adults (median: 0.01% [IQR 0.01–0.01%]; p= 0.004) (Figure 2e).

**Figure 2.**
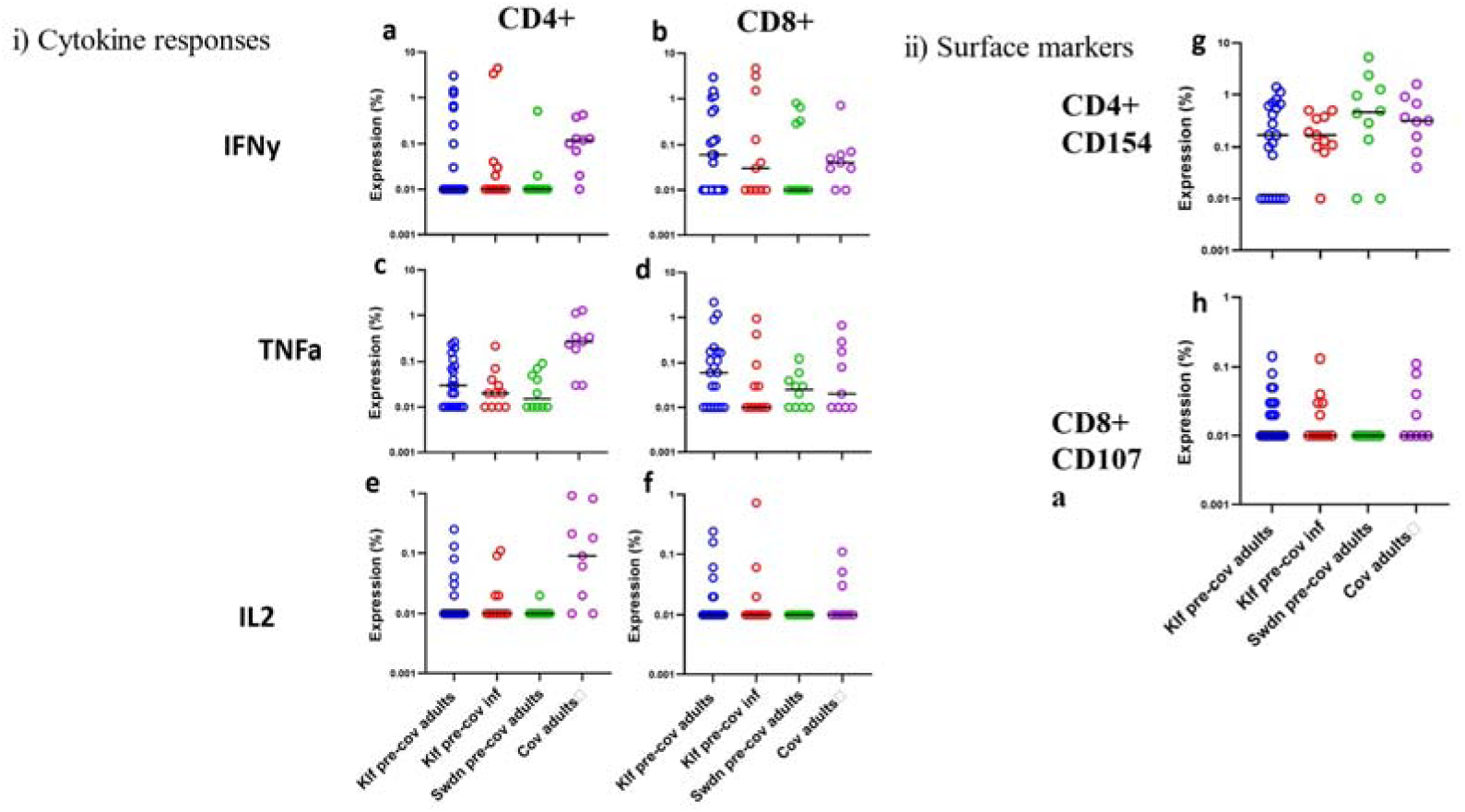
Proportions of T cells expressing cytokines and surface markers in response to SARS-CoV-2 Spike protein. i) Cytokine responses levels of **a** IFN-γ in CD4+ T cells, **b** IFN-γ in CD8+ T cells, **c** TNF in CD4+ T cells, **d** TNF in CD8+ T cells, **e** IL-2 in CD4+ T cells, **f** IL-2 in CD8+ T cells ii) Surface markers **g** CD154 on activated CD4+ T cells **h** CD107a on CD8+ T cells. Number of participants for: Kilifi pre-covid adults = 21, Kilifi pre-covid infants = 11, Sweden pre-covid adults = 10, Covid-19 adults = 9. Black line shows median.

Our analysis of the number of functional markers co-expressed by these T-cells showed that only a small proportion of individuals showed polyfunctionality prior to COVID-19. Individuals analysed post COVID-19 had better CD4 and CD8 T-cell polyfunctionality. Compared to Swedish pre-COVID-19 adults, a higher proportion of Kenyan pre-COVID-19 adults had polyfunctional CD4+ and CD8+ T-cells (Figure S4a, b). Collectively, these data suggest that the pre-COVID-19 CD4+ and CD8+T-cells from Swedish adults were less primed to secrete TNF-α and IFN-γ compared to those of the Kenyan pre-COVID-19 adults.

## Discussion

We compared the frequencies and prevalence of SARS-CoV-2 cross-reactive T-cell responses by measuring IFN-γ responses by ELISPOT and phenotypic and functional characterization by flow cytometry in pre-COVID-19 PBMC collected in Kenya, and in Sweden. We observed higher S1 specific and full spike frequencies, and a greater prevalence, of pre-pandemic SARS-CoV-2 spike peptide specific cross-reactive IFN-γ secreting cells measured by an ELISpot assay in Kenyan adults, compared to Swedish adults. Overall, these data suggest that Kenyan pre-COVID-19 adults had higher IFN-γ SFU responses to the S1 and spike regions but similar S2 responses compared to Swedish pre-COVID-19 adults. A larger proportion of Kenyan pre-COVID-19 adults also responded to SARS-CoV-2 spike peptides. Responses to S1-specific S2-specific, and to the full spike region increased following contact with COVID-19. These findings suggest a greater pre-pandemic exposure to S1-like epitopes in Kenya contributing to SARS-CoV-2 cross-reactive T-cell responses compared to Sweden.

We confirmed, using ICS assay, that the spike-peptide specific IFN-γ secreting cells included both CD4+ and CD8+ T-cells. Furthermore, we did not identify evidence that the Kenyan pre-COVID-19 adults had statistically significantly higher frequencies of pre-pandemic SARS-CoV-2 spike-specific TNF-α, or IFN-γ responses either in CD4+ T-cell responses or CD8+ T-cell responses compared to their Swedish counterparts. However, the ELISPOT assay is more sensitive than the ICS assay we used, and therefore the power to detect differences between populations is more limited in the ICS work.

The higher frequencies and prevalence of IFN-γ secreting cells among the Kenyan pre-pandemic adult-PBMCs compared to their Swedish counterparts is consistent with higher levels of pre-existing anti-SARS-CoV-2 cross protective immunity in SSA. These higher frequencies of pre-pandemic SARS-CoV-2 cross reactive cells may contribute to the lower prevalence of severe COVID-19 and deaths in Kenya relative to European countries like Sweden. This idea is consistent with previous observations of protection from severe COVID-19 by recent exposures to endemic hCoVs (27). Furthermore, mechanistic studies in mice demonstrated that hCoV-specific, SARS-CoV-2–cross-reactive T -cells contribute to SARS-CoV-2 immune responses upon infection and vaccination (17).

There is little evidence for a role for cross-reactive neutralising antibodies as these are rare, even where there is PCR evidence for recent hCoV infection (18). The mechanism by which SARS-CoV-2 cross reactive CD4+ T-cells lower COVID-19 severity may include various mechanisms such as the direct killing of virus infected cells, or through their ability to offer CD8 T cells help and protecting against infection, or both, thus reducing viral loads (28–30). Alternatively (or in addition), SARS-CoV-2 cross reactive memory CD4+ T-cells may provide the necessary costimulatory molecules for helping rapid and amnestic production of protective antibodies (17).

This study has a few limitations. First, for the ICS assays, we only had access to PBMC from 21 Kenyan pre-covid-19 adults,10 Kenyan pre-covid-19 infants, 10 Swedish pre-covid-19 adults and 9 Kenyan covid-19 adults, which reduced the statistical power of comparing responses among the groups. Second, we were only able to use spike peptides to stimulate PBMCs as the numbers of cells was limiting, it would have been more comprehensive to include other SARS-CoV-2 proteins.

## Conclusion

We report higher frequencies and prevalence of pre-COVID-19 SARS-CoV-2 spike cross-reactive IFN-γ secreting cells, some of which include CD8+ T -cells, among Kenyan compared to Swedish adults. This raises the possibility that higher levels of cross-protective cellular responses in SSA could explain, at least in part, why there was less severe morbidity and mortality on the continent relative to Europe, Asia and North America. Nonetheless, subsequent studies including relatively higher samples sizes of pre-COVID-19 PBMC from SSA and a few more countries from Europe and/or America are required to confirm these findings and extend the research into investigating the actual mechanism of the SARS-CoV-2 cross-protection. Importantly, such studies will provide new insights on how to develop novel cross-protective vaccines against similar pathogens, like coronaviruses.

## Data Availability

All data produced in the present study are in the process of being processed for uploading to the online Harvard Dataverse repository but are currently available upon request from the authors.

## Acknowledgments

We thank the field workers, laboratory staff and healthcare workers involved in the blood sampling. We appreciate all the study participants.

