## Supplementary material for "Pre-COVID-19 *ex vivo* cross-reactive IFN-γ cellular response to SARS-CoV-2 spike overlapping peptides is more robust among Kenyan compared to Swedish adults"

**Running Title**: Pre-COVID-19 cross-reactive IFN-γ responses to SARS-CoV-2 spike: Kenyan vs. Swedish adults

**Authors**

Perpetual Wanjiku^1^*, Benedict Orindi^1^, Jedidah Mwacharo^1^, James Chemweno^1^, Henry Karanja^1^, Barbara Kronsteiner^4,5^ Oscar Kai^1^, Daniel Wright^2^, Lynette Isabella Ochola-Oyier^1,3^, Christopher Sundling^6,7^, Susanna Dunachie^4,5^, George M Warimwe^1,3^, Anna Färnert^6,7^, Philip Bejon^1,8^, Francis M. Ndungu^1,3,6†^, Eunice Nduati^1,3†^.

**Affiliations**

1. Centre for Geographic Medicine Research (Coast), Kenya Medical Research Institute (KEMRI)-Wellcome Trust Research Programme, Kilifi, Kenya
2. Department of Pediatrics, University of Oxford, Oxford, United Kingdom
3. Centre for Tropical Medicine and Global Health, Nuffield Department of Medicine, University of Oxford, Oxford, United Kingdom
4. Centre for Global Health Research, Nuffield Department of Medicine, University of Oxford, Oxford, United Kingdom
5. Mahidol-Oxford Tropical Medicine Research Unit, Mahidol University, Thailand
6. Division of Infectious Diseases, Department of Medicine Solna, and Center for Molecular Medicine, Karolinska Institutet, Stockholm, Sweden
7. Department of Infectious Diseases, Karolinska University Hospital, Stockholm, Sweden
8. Modernising Medical Microbiology, Nuffield Department of Medicine, University of Oxford, Oxford, United Kingdom

†These authors contributed equally.

**Table S1. Participants tested for each peptide**

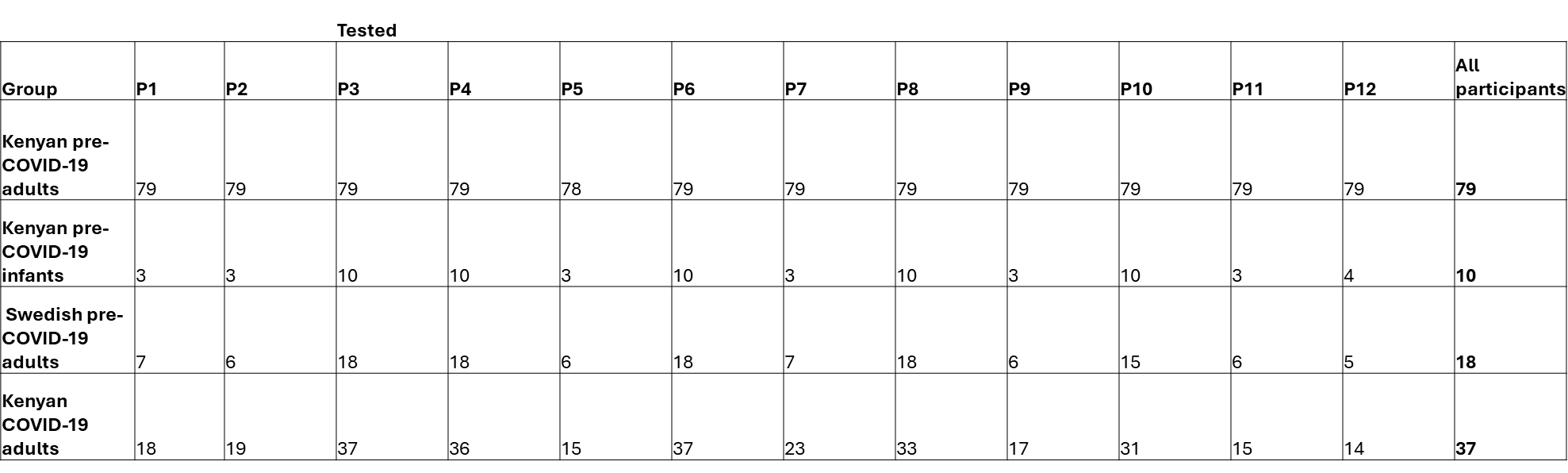

**Table S2. Participant demographic and clinical characteristics**†**.**

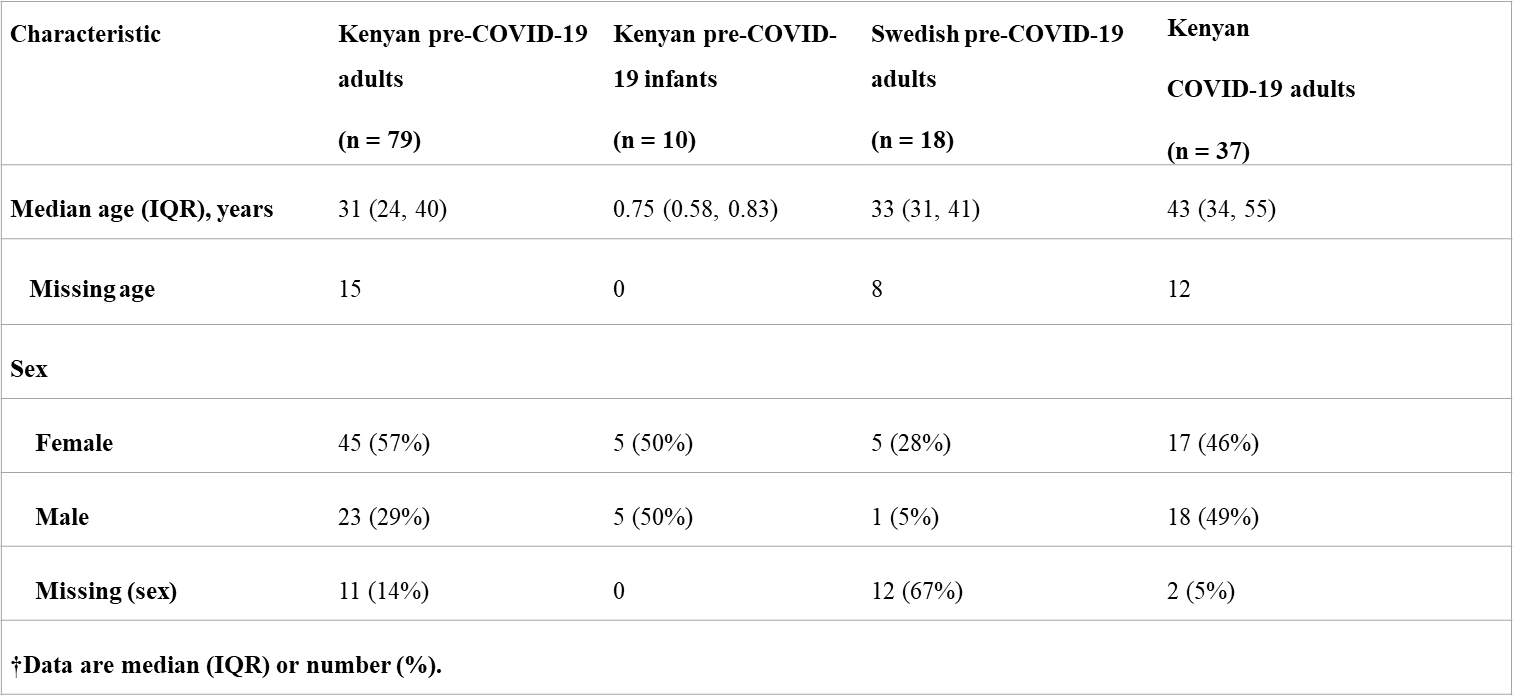

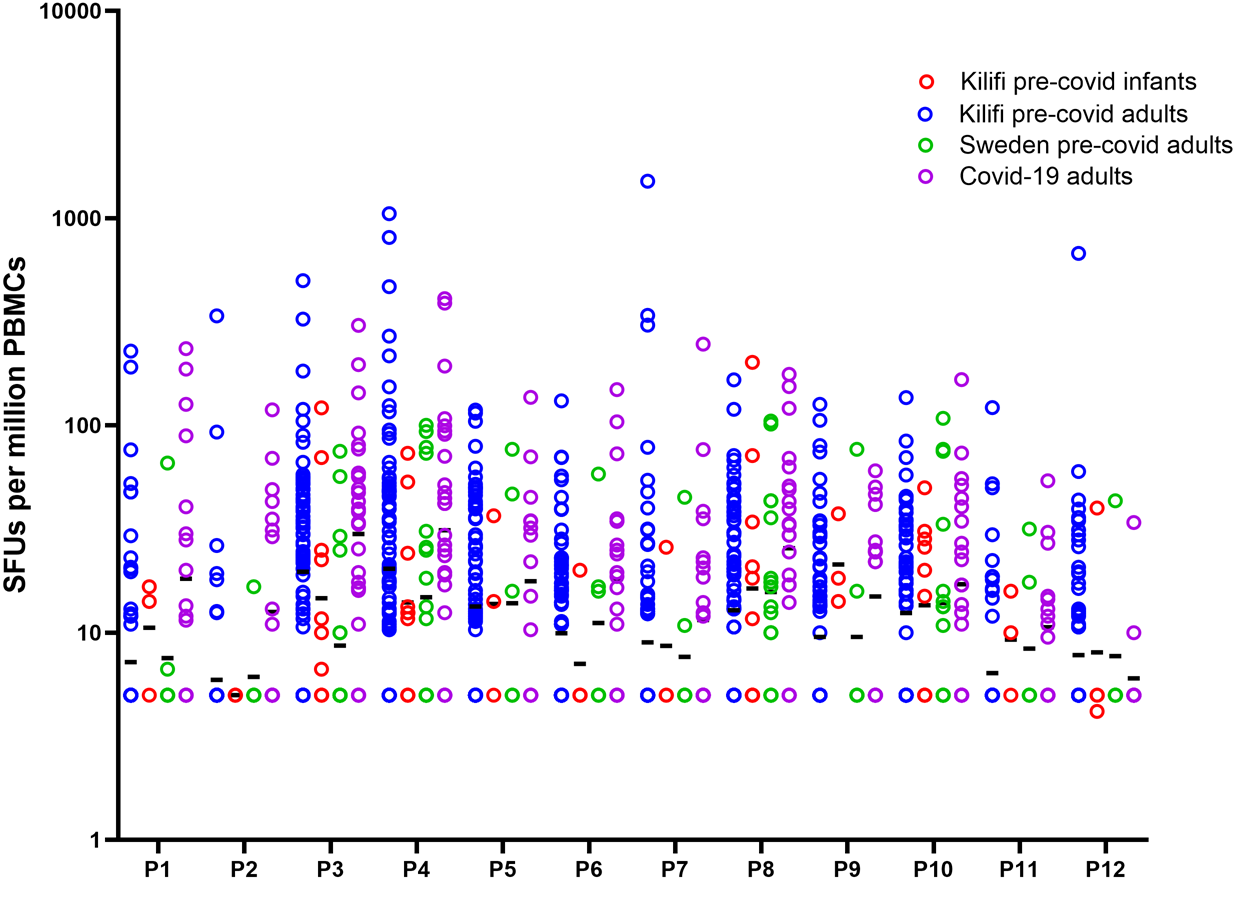

**Figure S1. Frequencies of IFN-gamma secreting cell responses to individual but overlapping SARS-CoV-2 Spike peptides pools (P1-P12).**
Black line shows Geometric mean

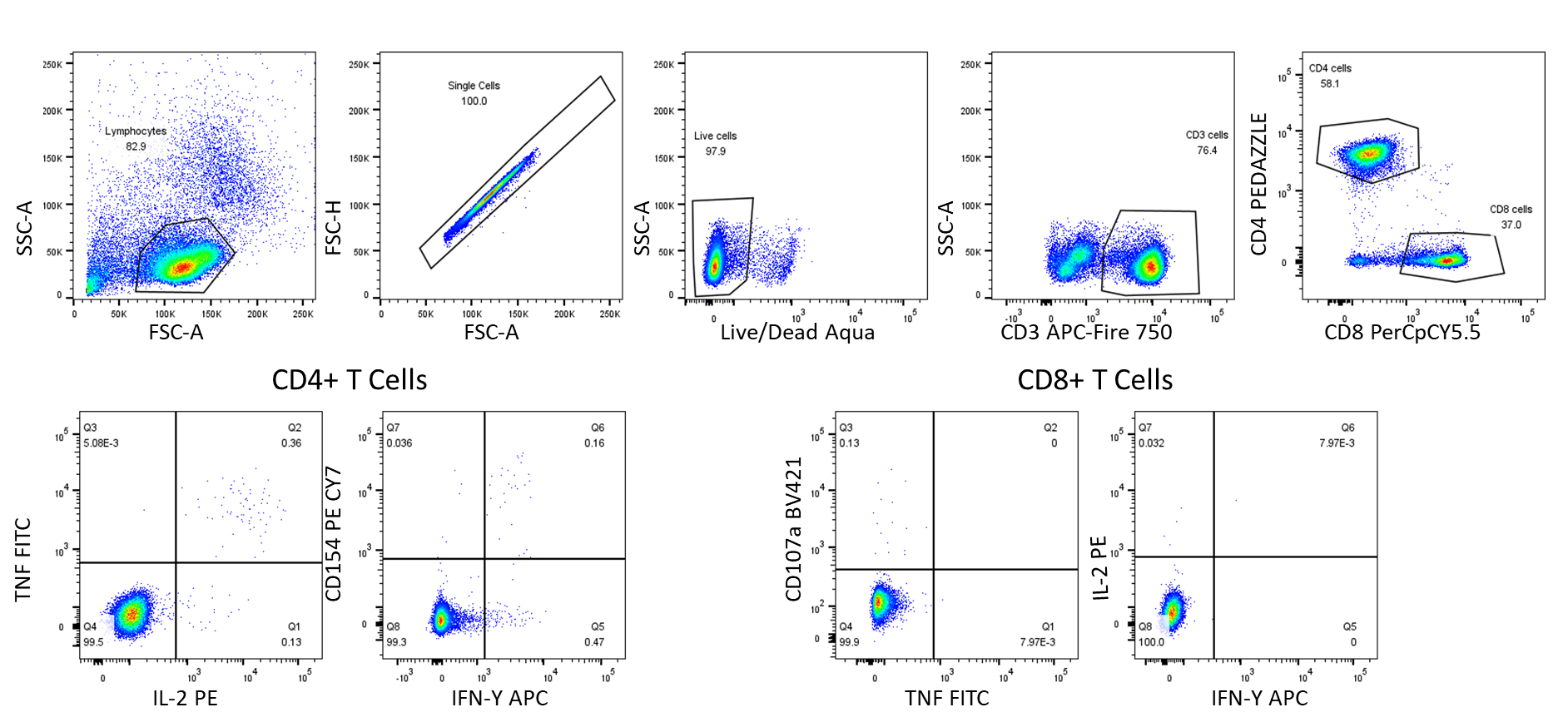

**Figure S2. ICS gating strategy.**

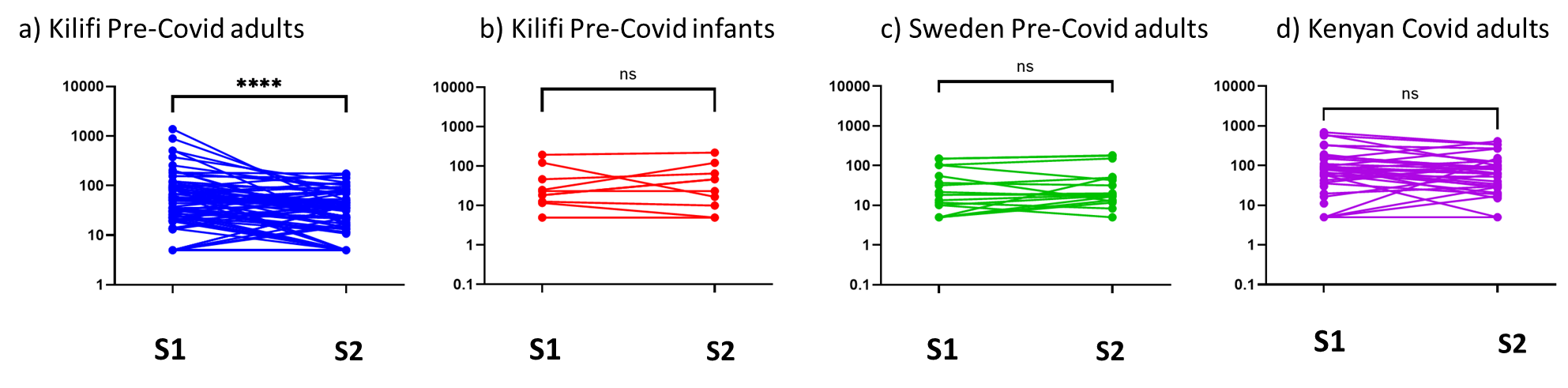

**Figure S3. Interindividual variability of IFN-gamma secreting cell frequencies between SARS-CoV-2 Spike regions S1 and S2.**

a) Kilifi pre-Covid adults, b) Kilifi Pre-Covid infants, c) Sweden Pre-Covid adults and d) Kenyan Covid adults. Comparisons were done using Wilcoxon matched pairs signed rank test. * P < 0.05, **P < 0.01, ***P < 0.001, ****P < 0.0001

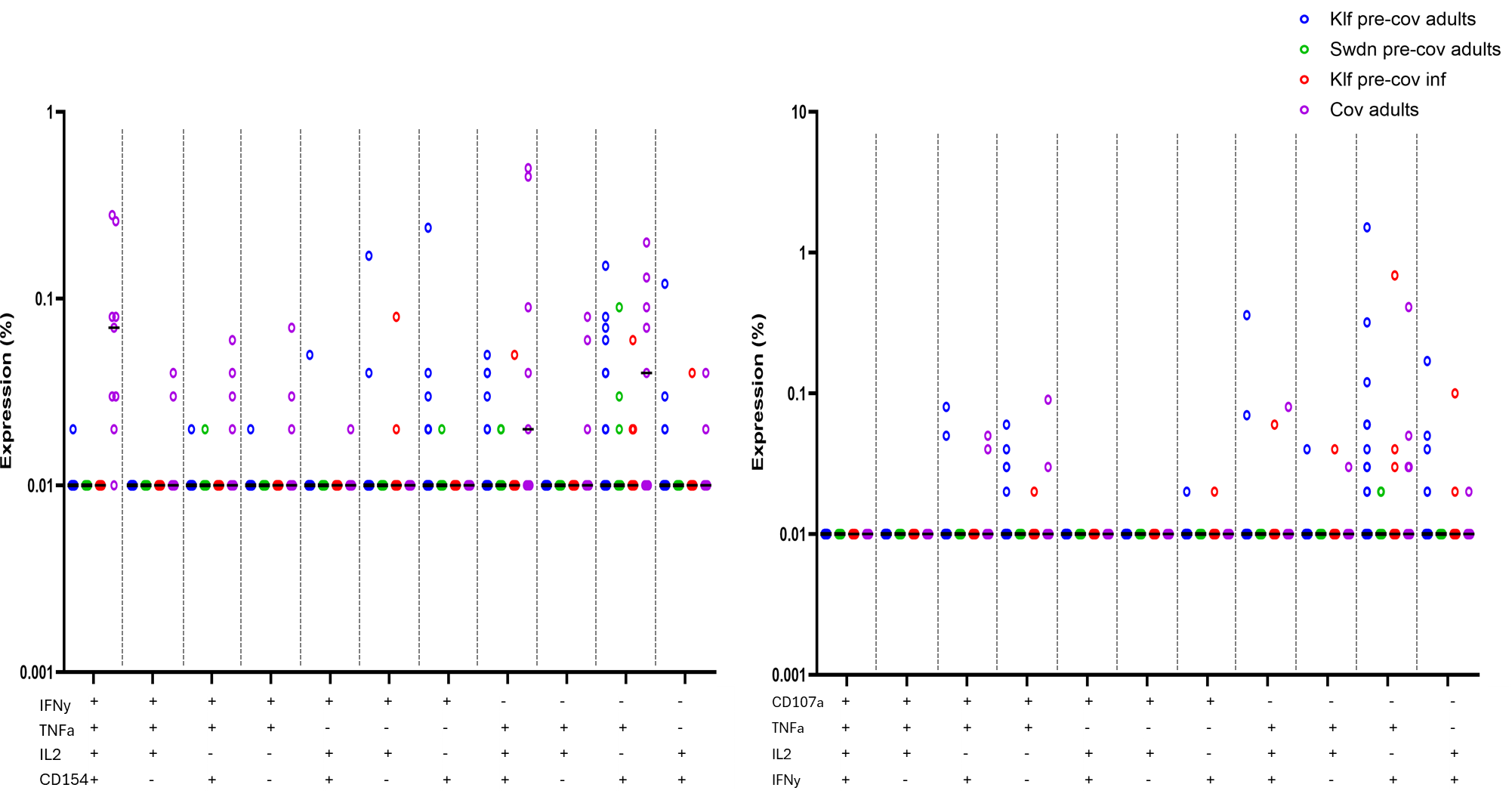

**Figure S4a. Polyfunctional expression profile of CD4+ T cells. Figure S4b. Polyfunctional expression profile of CD8 + T cells.**
